## Supplementary Note 1 for "Regional genetic correlations highlight relationships between neurodegenerative diseases and the immune system"

**Validation of significant regional genetic correlations with gene expression and protein levels, in AD and PD datasets without proxy cases**

We aimed at verifying that our results involving Alzheimer's disease (AD) and Parkinson's disease (PD) are not primarily driven by possible spurious effects of the inclusion of proxy cases in the GWAS samples. To do so, we performed regional genetic correlations with LAVA between the significant signals (FDR < 0.01) identified between AD or PD and gene expression or protein levels, using as input the AD or PD GWAS datasets without proxy cases (Blauwendraat et al., 2019; Kunkle et al., 2019). The aim was to compare results with or without the use of proxy cases for the AD and PD GWAS datasets.

### Genome-wide genetic correlations without proxy cases

We computed genome-wide genetic correlations between pairs of traits, including the AD and PD datasets without proxy cases (Figure 1). These GWASes exhibit a perfect global genetic correlation with the corresponding GWAS including proxy cases (AD: $r_g$ = 1.07 p-value = 9.86e-82; PD: $r_g$ = 1.04, p-value = 0). By using a Bonferroni-corrected p-value threshold (p-value < 0.0014), we identified the same significant correlations as in the main analysis, which used the AD and PD GWAS association results that included proxy cases. Previous studies have highlighted genome-wide genetic correlations among neurodegenerative diseases, such as between LBD and AD, for which we did not observe a significant correlation in our main analysis ($r_g$ = 0.33; p-value = 0.1324), and between AD and PD, for which we observed a nominal correlation when using the datasets with proxy cases ($r_g$ = 0.23; p-value = 0.0096). When using the AD dataset without proxy cases, we observed nominally significant genetic correlations between LBD and AD ($r_g$ = 0.69, p-value = 4.1e-03). For AD and PD, the correlation remained non-significant when using both the AD and PD datasets without proxies (rg = 0.12, p-value = 0.2).

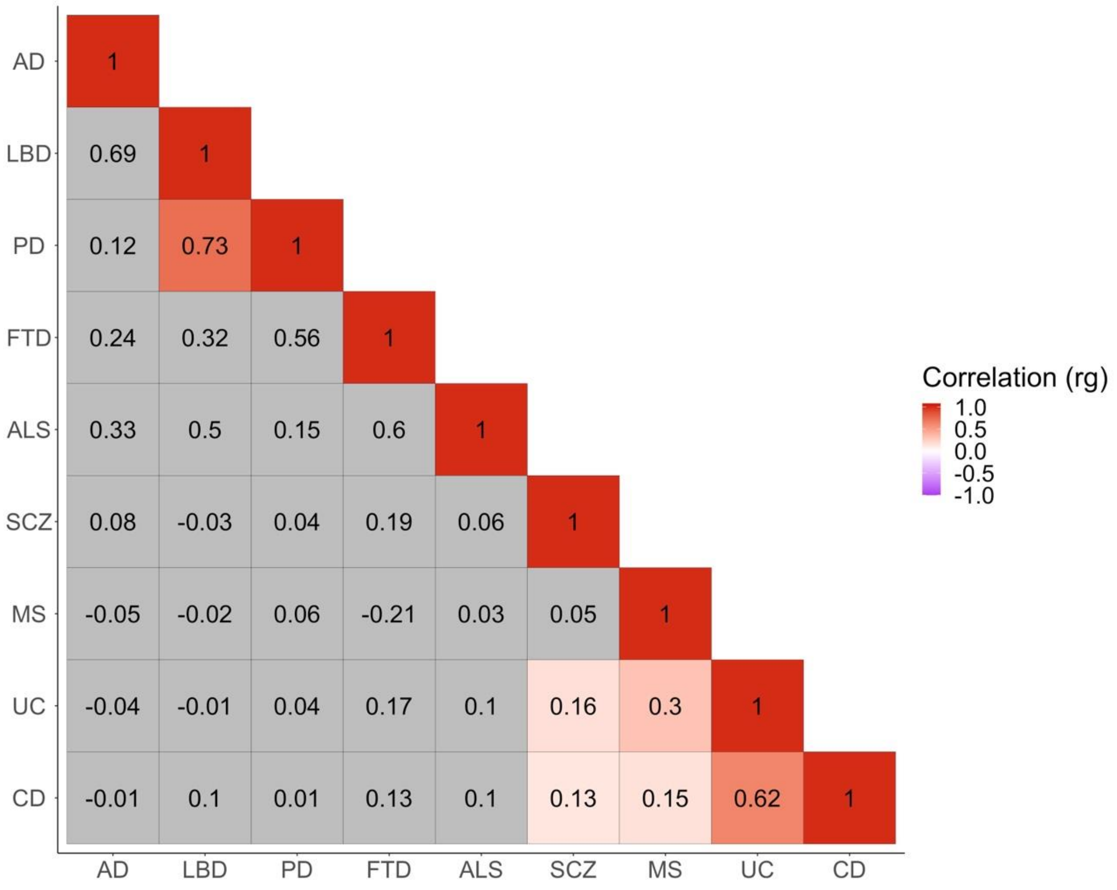


**Figure 1.** Genome-wide genetic correlations ($r_g$) across GWAS traits without AD or PD proxy cases. Significant positive correlations (Bonferroni corrected p-value < 0.0014) are highlighted in shades of red. AD = Alzheimer's disease; LBD = Lewy body dementia; PD = Parkinson's disease; FTD = frontotemporal dementia; ALS = amyotrophic lateral sclerosis; SCZ = schizophrenia; MS = multiple sclerosis; UC = ulcerative colitis; CD = Crohn's disease.

**Validation of significant correlations with gene expression levels**

In the primary analysis with proxy cases, we observed 30 significant correlations between AD and genes expressed across all seven cell types tested, encompassing 14 unique genes, including the HLA region. We reran LAVA across 21 out of the 30 significant AD correlations, given that nine of them did not have significant univariate signal in the AD GWAS without proxy cases (Table 1; Figure 2A). By using a Bonferroni p-value threshold for multiple testing (p-value < 0.05/30 tests performed for AD), we observed 12 significant correlations (p-value < 0.0016) and eight nominally significant correlations (p-value < 0.05) in the AD replication analysis for gene expression levels (Figure 2A). One significant correlation in the primary analysis did not replicate, implicating the expression of the gene *RP11_841O20_2* (Ensembl ID: ENSG00000258757) in CD4+ effector T cells ($r_g$ = 0.445; p-value = 0.125). However, the direction of effect was concordant in all tests (Spearman's correlation = 0.99; p-value = 2.2e-16) (Figure 2A).

In the primary correlation analysis between PD and gene expression levels, we observed 78 significant correlations across all seven cell types tested, encompassing 53 unique genes. We were able to perform the replication analysis for 10 of the primary significant correlations, given that 68 did not have sufficient univariate signal in the PD GWAS without proxy cases (Table 1; Figure 2B). We applied a Bonferroni p-value threshold accounting for 78 significant correlations observed in the primary analysis (p-value < 0.05/78 tests). There were five significant correlations (p-value < 0.0006) and five nominally significant correlations (p-value < 0.05) in the PD validation analysis for gene expression levels, all with the same direction of effect (Spearman's correlation = 0.80; p-value = 0.006) (Figure 2B).

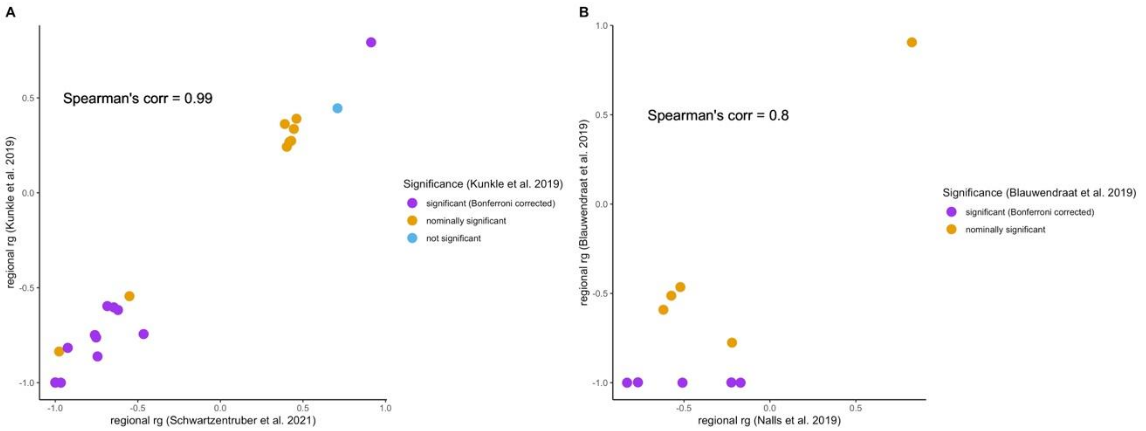


**Figure 2.** Scatter plots between the primary (i.e., with proxy cases) and replication (i.e., without proxy cases) regional genetic correlation analyses between (A) AD and gene expression levels, and (B) PD and gene expression levels.

**Validation of significant correlations with protein levels**

In the primary analysis with proxy cases, there were 10 significant correlations between AD and protein levels. From these, we performed the validation analysis at eight loci, given that two of them did not have sufficient univariate signal (Table 2; Figure 3A). After applying a Bonferroni p-value threshold for multiple testing (p-value < 0.05/10 tests), we observed seven significant correlations in the validation without proxies (p-value < 0.005) and one nominally significant correlation (p=value < 0.05). Additionally, the direction of effect was concordant between the primary and validation analyses (Spearman's correlation = 0.99; p-value = 1.589e-06) (Figure 3A).

In the primary correlations between PD and protein levels, there were 79 significant correlations. We performed the validation analysis only for six of the significant correlations that had sufficient univariate signal in the PD GWAS without proxy cases (Table 2; Figure 3B). By applying a Bonferroni p-value threshold to account for the number of significant correlations in the primary analysis (p-value < 0.05/79 tests), we observed three significant correlations (p-value < 0.0006) and three nominally significant correlations (p-value < 0.05), all of which had a concordant direction of effect (Spearman's correlation = 0.98; p-value = 0.0004) (Figure 3B).

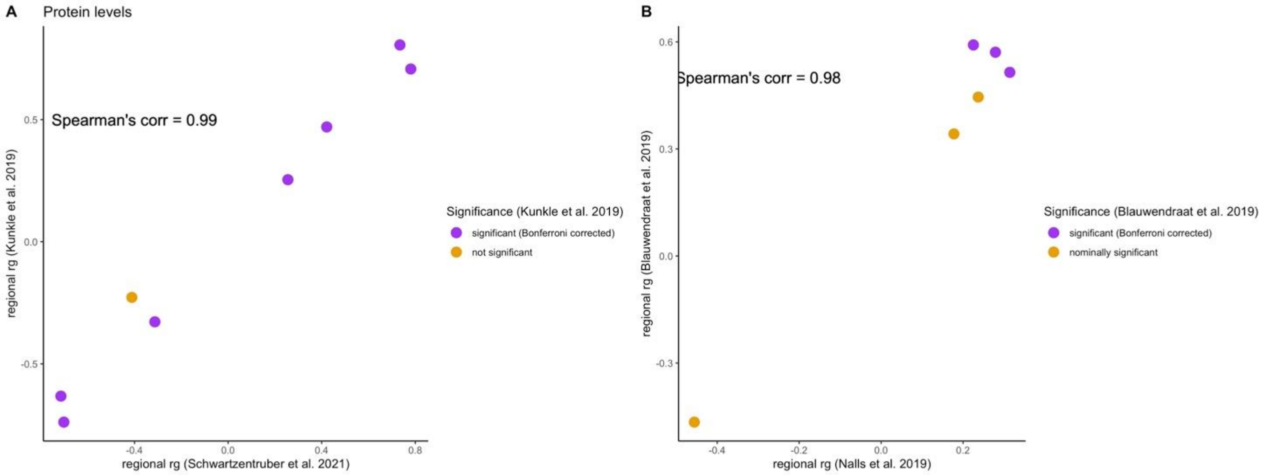


**Figure 3.** Scatter plots between the primary (i.e., with proxy cases) and validation (i.e., without proxy cases) regional genetic correlation analyses between (A) AD and protein levels, and (B) PD and protein levels.

**Table 1.** Validation of significant regional genetic correlations between AD or PD (using GWAS with proxy cases) and gene expression levels, using GWAS datasets without proxy cases.

| Disease | Gene Expressed | Cell type | Results (with proxy cases) | | Validation (without proxy cases) | |
|---|---|---|---|---|---|---|
| | | | $r_g$ | p-value | $r_g$ | p-value |
| AD | *BIN1* | BMem | 0.912 | 7.10E-06 | 0.792 | 7.94E-04 |
| AD | *BIN1* | CD4ET | 0.445 | 4.40E-04 | 0.336 | 0.015 |
| AD | *BIN1* | CD4NC | 0.402 | 2.82E-07 | 0.242 | 0.011 |
| AD | *BIN1* | CD8ET | 0.429 | 1.68E-07 | 0.274 | 4.93E-03 |
| AD | *BIN1* | CD8NC | 0.418 | 3.97E-05 | 0.268 | 0.020 |
| AD | *HLA_DRB1* | BIN | -0.465 | 1.12E-03 | -0.744 | 3.09E-04 |
| AD | *HLA_DQA1* | CD8ET | -1 | 6.70E-09 | -0.998 | 3.19E-04 |
| AD | *HLA_DQA1* | MonoC | -0.551 | 9.68E-04 | -0.544 | 0.022 |
| AD | *HLA_DQB1* | CD8ET | 0.39 | 3.07E-03 | 0.362 | 0.032 |
| AD | *HLA_DQB1* | MonoC | 0.46 | 8.82E-05 | 0.39 | 0.029 |
| AD | *HLA_DQA2* | BIN | -0.685 | 2.22E-10 | -0.597 | 1.98E-04 |
| AD | *HLA_DQA2* | BMem | -0.62 | 1.34E-06 | -0.617 | 2.11E-04 |
| AD | *HLA_DQA2* | CD8ET | -0.645 | 1.71E-12 | -0.603 | 2.92E-06 |
| AD | *HLA_DQA2* | CD8NC | -0.744 | 4.54E-09 | -0.862 | 2.72E-05 |
| AD | *HLA_DQA2* | MonoC | -0.966 | 1.41E-10 | -1 | 7.72E-06 |
| AD | *GATS* | BIN | -0.977 | 3.39E-06 | -0.836 | 0.011 |
| AD | *EPHA1_AS1* | MonoC | -1 | 3.06E-09 | -1 | 2.50E-07 |
| AD | *FNBP4* | BMem | -0.924 | 3.32E-04 | -0.816 | 6.49E-04 |
| AD | *FNBP4* | CD8ET | -0.753 | 2.18E-03 | -0.762 | 8.89E-04 |

| | | | | | | |
|---|---|---|---|---|---|---|
| AD | *FNBP4* | CD8NC | -0.76 | 2.03E-03 | -0.749 | 1.58E-03 |
| AD | *RP11_841O20_2* | CD4ET | 0.709 | 1.82E-03 | 0.445 | 0.125 |
| PD | *RAB7L1* | CD4NC | 0.826 | 1.06E-03 | 0.905 | 8.22E-04 |
| PD | *HLA_DRB5* | MonoC | -0.221 | 6.91E-04 | -0.776 | 2.44E-03 |
| PD | *HMBOX1* | BIN | -0.574 | 3.19E-04 | -0.513 | 1.00E-03 |
| PD | *HMBOX1* | BMem | -0.62 | 8.75E-04 | -0.592 | 9.93E-04 |
| PD | *HMBOX1* | CD8NC | -0.521 | 3.17E-03 | -0.465 | 6.33E-03 |
| PD | *CRHR1* | CD8NC | -0.509 | 4.29E-04 | -1 | 3.18E-06 |
| PD | *KANSL1_AS1* | BIN | -0.225 | 1.24E-05 | -0.999 | 2.44E-11 |
| PD | *KANSL1_AS1* | BMem | -0.171 | 1.73E-03 | -1 | 5.51E-11 |
| PD | *KANSL1_AS1* | CD8ET | -0.831 | 1.34E-39 | -1 | 6.51E-08 |
| PD | *KANSL1_AS1* | CD8NC | -0.768 | 1.13E-25 | -0.998 | 1.73E-09 |

BIN = naïve B cells; BMem = Memory B cells; CD4ET = CD4+ effector memory T cells; CD4NC = CD4+ naïve T cells; CD8ET = CD8+ effector memory T cells; CD8NC = CD8+ naïve T cells; MonoC = classical monocytes.

**Table 2.** Validation of significant regional genetic correlations between AD or PD (using GWAS with proxy cases) and protein levels, using GWAS datasets without proxy cases.

| Disease | Protein gene symbol | Results (with proxy cases) | | Validation (without proxy cases) | |
|---|---|---|---|---|---|
| | | $r_g$ | p-value | $r_g$ | p-value |
| AD | *CR1* | 0.734 | 6.00E-20 | 0.806 | 4.00E-15 |
| AD | *BIN1* | 0.781 | 2.90E-19 | 0.707 | 3.88E-12 |
| AD | *C2* | -0.702 | 1.09E-07 | -0.738 | 1.51E-05 |
| AD | *ATF6B* | -0.411 | 1.37E-03 | -0.228 | 0.149 |
| AD | *TREM2* | -0.714 | 6.58E-07 | -0.632 | 1.89E-04 |
| AD | *PLCG2* | -0.313 | 3.03E-04 | -0.328 | 2.59E-03 |
| AD | *APOE* | 0.421 | 1.08E-05 | 0.47 | 1.66E-06 |
| AD | *APOC1* | 0.255 | 3.51E-09 | 0.254 | 2.51E-08 |
| PD | *FCGR2A* | 0.237 | 6.02E-13 | 0.445 | 8.46E-04 |
| PD | *CNTN2* | 0.178 | 4.57E-04 | 0.342 | 6.83E-03 |
| PD | *BST1* | 0.314 | 2.87E-07 | 0.515 | 3.68E-04 |
| PD | *TREML2* | 0.279 | 1.72E-04 | 0.571 | 7.65E-05 |
| PD | *PCBD1* | 0.225 | 3.17E-04 | 0.591 | 1.34E-04 |
| PD | *SIGLEC15* | -0.456 | 3.97E-05 | -0.466 | 0.0176 |
