## Supplementary Tables for "Regional genetic correlations highlight relationships between neurodegenerative diseases and the immune system"

**Supplementary Table 1.** Overview of the GWAS summary statistics used in the present study.

| GWAS (acronym) | N cases | N controls | N variants | Chromosomes | Genome build | LDSC observed SNP $h^2$ (Z-score) | Reference | URL |
|---|---|---|---|---|---|---|---|---|
| Alzheimer's disease (AD) | 75,671 (52,791 proxies) | 397,844 | 10,687,077 | 1-22 | GRCh37 | 1.06 % (1.860) | Schwartzentruber *et al.* 2021 | http://ftp.ebi.ac.uk/pub/databases/gwas/summary_statistics/GCST90012001-GCST90013000/GCST90012877/ |
| Amyotrophic lateral sclerosis (ALS) | 27,205 | 110,881 | 10,461,755 | 1-22 | GRCh37 | 3.82 % (8.489) | Van Rheenen *et al.* 2021 | https://www.projectmine.com/research/download-data/ |
| Crohn's disease (CD) | 12,194 | 28,072 | 9,550,617 | 1-22 | GRCh37 | 44.59 % (8.847) | de Lange *et al.* 2017 | http://ftp.ebi.ac.uk/pub/databases/gwas/summary_statistics/GCST004001-GCST005000/GCST004132/ |
| Frontotemporal dementia (FTD) | 2,154 | 4,308 | 6,026,384 | 1-22 | GRCh37 | 8.08 % (1.135) | Ferrari *et al.* 2014 | https://ifgcsite.wordpress.com/data-access/ |
| Lewy body dementia (LBD) | 2,591 | 4,027 | 7,827,747 | 1-22 | GRCh38 | 14.81 % (1.806) | Chia *et al.* 2021 | http://ftp.ebi.ac.uk/pub/databases/gwas/summary_statistics/GCST90001001-GCST90002000/GCST90001390/ |

| | | | | | | | | |
|---|---|---|---|---|---|---|---|---|
| Multiple sclerosis (MS) | 14,802 | 26,703 | 8,244,101 | 1-22, X, Y | GRCh37 | 29.33 % (10.863) | IMSGC 2019 | Requested access through https://imsgc.net/?page_id=31 |
| Parkinson's disease (PD) | 33,674 (18,618 proxies) | 449,056 | 12,076,399 | 1-22 | GRCh37 | 1.62 % (10.125) | Nalls *et al.* 2019 | https://www.pdgenetics.org/resources |
| Schizophrenia (SCZ) | 40,675 | 64,643 | 8,064,799 | 1-22, X | GRCh37 | 42.17 % (28.113) | Pardiñas *et al.* 2018 | https://pgc.unc.edu/for-researchers/download-results/ |
| Ulcerative colitis (UC) | 12,366 | 33,609 | 9,567,780 | 1-22 | GRCh37 | 23.97 % (9.255) | de Lange *et al.* 2017 | http://ftp.ebi.ac.uk/pub/databases/gwas/summary_statistics/GCST004001-GCST005000/GCST004133/ |

**Supplementary Table 2.** Genome-wide genetic correlations between pairs of diseases evaluated in the study estimated with LD Score Regression. $r_g$ = genetic correlation; SE = standard error. ns = non-significant correlations; • = nominal correlations (p-value < 0.05); ** = Bonferroni-corrected significant correlations (p-value < 0.0014).

| GWAS trait 1 | GWAS trait 2 | $r_g$ | SE | p-value | Significance |
|---|---|---|---|---|---|
| AD | ALS | 0.19 | 0.089 | 0.0359 | • |
| AD | CD | -0.13 | 0.053 | 0.0125 | • |
| AD | FTD | 0.29 | 0.291 | 0.3126 | ns |
| AD | LBD | 0.33 | 0.221 | 0.1324 | ns |
| AD | MS | -0.06 | 0.059 | 0.2899 | ns |
| AD | PD | 0.23 | 0.089 | 0.0096 | • |
| AD | SCZ | 0.03 | 0.038 | 0.4934 | ns |
| AD | UC | -0.08 | 0.066 | 0.2038 | ns |

| | | | | | |
|---|---|---|---|---|---|
| ALS | CD | 0.1 | 0.051 | 0.0618 | ns |
| ALS | FTD | 0.6 | 0.430 | 0.164 | ns |
| ALS | LBD | 0.5 | 0.208 | 0.0168 | • |
| ALS | MS | 0.03 | 0.060 | 0.6411 | ns |
| ALS | PD | 0.14 | 0.065 | 0.0334 | • |
| ALS | SCZ | 0.06 | 0.036 | 0.0722 | ns |
| ALS | UC | 0.1 | 0.055 | 0.0634 | ns |
| CD | FTD | 0.13 | 0.144 | 0.3724 | ns |
| CD | LBD | 0.1 | 0.109 | 0.3797 | ns |
| CD | MS | 0.15 | 0.047 | 0.001 | ** |
| CD | PD | 0 | 0.040 | 0.998 | ns |
| CD | SCZ | 0.13 | 0.024 | 1.68E-07 | ** |
| CD | UC | 0.62 | 0.033 | 2.43E-80 | ** |
| FTD | LBD | 0.32 | 0.537 | 0.5546 | ns |
| FTD | MS | -0.21 | 0.193 | 0.2745 | ns |
| FTD | PD | 0.48 | 0.285 | 0.0926 | ns |
| FTD | SCZ | 0.19 | 0.153 | 0.2196 | ns |
| FTD | UC | 0.17 | 0.184 | 0.3589 | ns |
| LBD | AD | 0.33 | 0.221 | 0.1324 | ns |
| LBD | MS | -0.02 | 0.113 | 0.8685 | ns |
| LBD | PD | 0.65 | 0.196 | 0.001 | ** |
| LBD | SCZ | -0.03 | 0.075 | 0.6509 | ns |
| LBD | UC | -0.01 | 0.130 | 0.9129 | ns |
| MS | PD | 0.03 | 0.045 | 0.472 | ns |
| MS | SCZ | 0.05 | 0.029 | 0.1197 | ns |
| MS | UC | 0.3 | 0.045 | 1.45E-11 | ** |
| PD | SCZ | 0.02 | 0.030 | 0.5272 | ns |
| PD | UC | 0.05 | 0.049 | 0.2795 | ns |
| SCZ | UC | 0.16 | 0.028 | 1.90E-08 | ** |

**Supplementary Table 3.** Number of genome-wide significantly expressed genes per cell type from the OneK1K single-cell eQTL data.

| Cell Type | N significantly expressed genes |
|---|---|
| Classical monocytes | 196 |
| CD4+ naïve T cells | 2,089 |
| CD4+ effector memory T cells | 543 |
| CD8+ naïve T cells | 930 |
| CD8+ effector memory T cells | 1,018 |
| Naïve B cells | 502 |
| Memory B cells | 383 |

**Supplementary Table 4.** Colocalization results between diseases and expressed genes across immune cell types for disease-cell type pairs with Posterior Probability, PP H4 ≥ 0.8, indicating evidence of a shared causal signal.

| GWAS trait | Cell type | Chr | Gene expressed | N SNPs | PP (H4) |
|---|---|---|---|---|---|
| UC | Effector memory CD8+ T cells | 1 | *TNFRSF14* | 323 | 0.98 |
| SCZ | Naïve/central memory CD8+ T cells | 1 | *CTSS* | 299 | 0.96 |
| CD | Naïve/central memory CD8+ T cells | 1 | *SCAMP3* | 161 | 0.84 |
| MS | Immature and naïve B cells | 1 | *FCRL3* | 456 | 0.94 |
| MS | Effector memory CD8+ T cells | 1 | *FCRL3* | 456 | 0.82 |
| MS | Immature and naïve B cells | 1 | *RGS1* | 332 | 0.93 |
| PD | Naïve/central memory CD4+ T cells | 1 | *RAB7L1* | 311 | 0.93 |
| SCZ | Naïve/central memory CD4+ T cells | 2 | *FOXN2* | 504 | 0.99 |
| SCZ | Effector memory CD8+ T cells | 2 | *FOXN2* | 504 | 0.99 |
| SCZ | Naïve/central memory CD8+ T cells | 2 | *FOXN2* | 504 | 0.99 |
| MS | Naïve/central memory CD4+ T cells | 2 | *PLEK* | 557 | 0.88 |
| SCZ | Effector memory CD8+ T cells | 3 | *PPM1M* | 226 | 0.99 |
| SCZ | Naïve/central memory CD8+ T cells | 3 | *PPM1M* | 226 | 0.99 |
| MS | Immature and naïve B cells | 3 | *EAF2* | 593 | 0.92 |

| | | | | | |
|---|---|---|---|---|---|
| MS | Memory B cells | 3 | *EAF2* | 593 | 0.92 |
| SCZ | Effector memory CD8+ T cells | 5 | *EMB* | 502 | 0.89 |
| CD | Immature and naïve B cells | 5 | *ERAP2* | 766 | 0.87 |
| CD | Memory B cells | 5 | *ERAP2* | 766 | 0.88 |
| CD | Effector memory CD4+ T cells | 5 | *ERAP2* | 766 | 0.86 |
| CD | Effector memory CD8+ T cells | 5 | *ERAP2* | 766 | 0.86 |
| CD | Naïve/central memory CD8+ T cells | 5 | *ERAP2* | 766 | 0.85 |
| UC | Naïve/central memory CD8+ T cells | 5 | *RGS14* | 194 | 0.99 |
| MS | Effector memory CD8+ T cells | 6 | *HIST1H1D* | 419 | 0.93 |
| MS | Immature and naïve B cells | 6 | *AHI1* | 332 | 0.99 |
| MS | Memory B cells | 6 | *AHI1* | 332 | 0.99 |
| MS | Effector memory CD4+ T cells | 6 | *AHI1* | 332 | 0.99 |
| MS | Naïve/central memory CD4+ T cells | 6 | *AHI1* | 332 | 0.99 |
| MS | Effector memory CD8+ T cells | 6 | *AHI1* | 332 | 0.99 |
| MS | Naïve/central memory CD8+ T cells | 6 | *AHI1* | 332 | 0.99 |
| SCZ | Effector memory CD8+ T cells | 7 | *MAD1L1* | 1300 | 0.91 |
| UC | Memory B cells | 7 | *LAMB1* | 496 | 0.97 |
| MS | Immature and naïve B cells | 8 | *ZC2HC1A* | 442 | 0.94 |
| MS | Memory B cells | 8 | *ZC2HC1A* | 442 | 0.94 |
| MS | Naïve/central memory CD4+ T cells | 8 | *ZC2HC1A* | 442 | 0.93 |
| CD | Effector memory CD4+ T cells | 11 | *IFITM2* | 612 | 1.00 |
| MS | Monocytes | 11 | *IFITM3* | 610 | 1.00 |
| AD | Memory B cells | 11 | *FNBP4* | 319 | 0.91 |
| CD | Effector memory CD4+ T cells | 11 | *TMEM258* | 341 | 0.93 |
| SCZ | Naïve/central memory CD8+ T cells | 11 | *RNASEH2C* | 373 | 0.82 |
| MS | Immature and naïve B cells | 12 | *CLEC2D* | 700 | 0.98 |
| MS | Effector memory CD8+ T cells | 12 | *CLECL1* | 660 | 0.97 |
| MS | Naïve/central memory CD8+ T cells | 12 | *CLECL1* | 660 | 0.93 |

| | | | | | |
|---|---|---|---|---|---|
| MS | Effector memory CD4+ T cells | 12 | *METTL21B* | 217 | 0.93 |
| MS | Naïve/central memory CD8+ T cells | 12 | *METTL21B* | 217 | 0.93 |
| CD | Immature and naïve B cells | 16 | *TUFM* | 142 | 0.97 |
| CD | Memory B cells | 16 | *TUFM* | 142 | 0.93 |
| CD | Effector memory CD4+ T cells | 16 | *TUFM* | 142 | 0.96 |
| CD | Effector memory CD8+ T cells | 16 | *TUFM* | 142 | 0.94 |
| CD | Naïve/central memory CD8+ T cells | 16 | *TUFM* | 142 | 0.97 |
| CD | Monocytes | 16 | *TUFM* | 142 | 0.90 |
| UC | Effector memory CD8+ T cells | 16 | *BRD7* | 427 | 0.87 |
| SCZ | Naïve/central memory CD8+ T cells | 16 | *NUTF2* | 134 | 0.96 |
| AD | Immature and naïve B cells | 17 | *SCIMP* | 437 | 0.99 |
| AD | Memory B cells | 17 | *SCIMP* | 437 | 0.86 |
| CD | Effector memory CD4+ T cells | 17 | *ORMDL3* | 380 | 0.95 |
| UC | Effector memory CD4+ T cells | 17 | *ORMDL3* | 380 | 0.95 |
| PD | Immature and naïve B cells | 17 | *KANSL1_AS1* | 213 | 0.88 |
| PD | Memory B cells | 17 | *KANSL1_AS1* | 213 | 0.87 |
| PD | Naïve/central memory CD4+ T cells | 17 | *KANSL1_AS1* | 213 | 0.88 |
| PD | Naïve/central memory CD8+ T cells | 17 | *KANSL1_AS1* | 213 | 0.87 |
| CD | Naïve/central memory CD4+ T cells | 19 | *AKAP8* | 159 | 0.83 |
| CD | Naïve/central memory CD8+ T cells | 19 | *AKAP8* | 159 | 0.90 |
| MS | Naïve/central memory CD4+ T cells | 19 | *MPV17L2* | 477 | 0.83 |
| UC | Naïve/central memory CD4+ T cells | 19 | *PTGIR* | 399 | 0.96 |
| UC | Naïve/central memory CD8+ T cells | 19 | *PTGIR* | 399 | 0.98 |
| UC | Naïve/central memory CD4+ T cells | 19 | *GNG8* | 361 | 0.97 |
| MS | Immature and naïve B cells | 19 | *CD37* | 399 | 0.99 |
| MS | Memory B cells | 19 | *CD37* | 399 | 0.97 |
| MS | Effector memory CD4+ T cells | 19 | *CD37* | 399 | 0.98 |
| MS | Naïve/central memory CD4+ T cells | 19 | *CD37* | 399 | 0.97 |

| | | | | | |
|---|---|---|---|---|---|
| MS | Effector memory CD8+ T cells | 19 | *CD37* | 399 | 0.97 |
| SCZ | Naïve/central memory CD4+ T cells | 19 | *IRF3* | 218 | 0.95 |
| CD | Naïve/central memory CD8+ T cells | 20 | *EDN3* | 304 | 0.98 |
| CD | Effector memory CD8+ T cells | 22 | *UBE2L3* | 262 | 0.95 |
| UC | Immature and naïve B cells | 22 | *SYNGR1* | 444 | 0.83 |
| SCZ | Monocytes | 22 | *SMDT1* | 354 | 0.88 |
| UC | Memory B cells | 22 | *PIM3* | 407 | 0.97 |
| MS | Memory B cells | 22 | *TYMP* | 489 | 0.99 |
| PD | Effector memory CD8+ T cells | 22 | *ARSA* | 435 | 0.88 |

AD = Alzheimer's disease; CD = Crohn's disease; PD = Parkinson's disease; UC = Ulcerative colitis; MS = Multiple sclerosis; SCZ = Schizophrenia

**Supplementary Table 5.** Regional genetic correlations between diseases and gene expression levels, for which the implicated loci did not harbour genome-wide significant GWAS variants (p-value < 5E-8).

| Gene expressed | Chr | N SNPs in model | GWAS trait | Cell type | Local $r_g$ | p-value | SNP with most significant GWAS p-value | Most significant GWAS p-value |
|---|---|---|---|---|---|---|---|---|
| *PARK7* | 1 | 49 | CD | CD8NC | -0.732 | 3.753E-03 | rs35087108 | 1.36E-07 |
| *RP11_108M9_4* | 1 | 40 | PD | CD8ET | -0.622 | 2.728E-03 | rs190628768 | 0.0002907 |
| *AK5* | 1 | 28 | PD | CD8ET | -0.668 | 1.073E-03 | rs17149959 | 7.09E-05 |
| *CTSS* | 1 | 8 | SCZ | CD8NC | 1.000 | 1.945E-04 | rs114845445 | 7.01E-08 |
| *FOXN2* | 2 | 82 | SCZ | CD4NC | -0.682 | 5.065E-04 | rs60710459 | 1.62E-07 |
| *FOXN2* | 2 | 4 | SCZ | CD8ET | -1.000 | 1.312E-04 | rs60710459 | 1.62E-07 |
| *FOXN2* | 2 | 5 | SCZ | CD8NC | -1.000 | 2.113E-04 | rs60710459 | 1.62E-07 |
| *MAL* | 2 | 55 | PD | CD4NC | -0.545 | 7.777E-04 | rs61771834 | 0.0003929 |
| *SLC41A3* | 3 | 148 | PD | CD4NC | 0.534 | 1.846E-03 | rs6470265 | 0.0005132 |
| *SPINK2* | 4 | 56 | PD | CD4NC | 0.272 | 6.256E-04 | rs73129300 | 1.84E-05 |

| | | | | | | | | |
|---|---|---|---|---|---|---|---|---|
| *PWWP2A* | 5 | 215 | PD | CD4NC | 0.369 | 4.399E-04 | rs75861893 | 0.0002433 |
| *DSP* | 6 | 96 | PD | BIN | 0.319 | 2.285E-04 | rs13359813 | 9.61E-05 |
| *MBOAT1* | 6 | 604 | PD | CD4NC | 0.333 | 2.732E-03 | rs11646153 | 8.89E-05 |
| *HIST1H1D* | 6 | 66 | MS | CD8ET | -0.767 | 3.087E-03 | rs1997768 | 9.06E-07 |
| *HLA_B* | 6 | 204 | AD | BIN | 0.590 | 1.685E-04 | rs6923313 | 8.64E-08 |
| *HLA_B* | 6 | 18 | PD | BIN | 0.539 | 7.385E-04 | rs191516377 | 0.0002219 |
| *HLA_B* | 6 | 415 | AD | CD4ET | 0.604 | 1.159E-05 | rs6923313 | 8.64E-08 |
| *HLA_B* | 6 | 35 | PD | CD4ET | 0.400 | 2.821E-03 | rs191516377 | 0.0002219 |
| *HLA_B* | 6 | 531 | AD | CD8ET | 0.561 | 3.100E-05 | rs6923313 | 8.64E-08 |
| *HLA_B* | 6 | 51 | PD | CD8ET | 0.453 | 8.857E-04 | rs191516377 | 0.0002219 |
| *HLA_B* | 6 | 26 | PD | CD8NC | 0.470 | 4.618E-07 | rs191516377 | 0.0002219 |
| *HLA_B* | 6 | 269 | AD | MonoC | 0.759 | 1.022E-06 | rs6923313 | 8.64E-08 |
| *HLA_DRB5* | 6 | 67 | ALS | BIN | -0.543 | 8.042E-04 | rs9268833 | 1.97E-07 |
| *HLA_DRB5* | 6 | 95 | ALS | BMem | -0.578 | 6.026E-04 | rs9268833 | 1.97E-07 |
| *HLA_DRB5* | 6 | 101 | ALS | CD8ET | -0.584 | 2.278E-06 | rs9268833 | 1.97E-07 |
| *CCDC167* | 6 | 138 | PD | CD4NC | -0.224 | 1.064E-05 | rs187517328 | 9.89E-05 |
| *RP1_8B1_4* | 6 | 150 | PD | CD8ET | 0.484 | 1.325E-03 | rs12580303 | 0.0003288 |
| *SFT2D1* | 6 | 41 | PD | CD8NC | -0.487 | 6.885E-04 | rs1577548 | 1.69E-05 |
| *CCZ1B* | 7 | 127 | PD | CD8ET | -0.317 | 9.332E-06 | rs56145568 | 0.0001754 |
| *HMBOX1* | 8 | 171 | PD | BIN | -0.574 | 3.194E-04 | rs6808178 | 7.20E-07 |
| *HMBOX1* | 8 | 177 | PD | BMem | -0.620 | 8.747E-04 | rs6808178 | 7.20E-07 |
| *HMBOX1* | 8 | 185 | PD | CD8NC | -0.521 | 3.169E-03 | rs6808178 | 7.20E-07 |
| *PKIA* | 8 | 116 | PD | CD4NC | -0.670 | 7.061E-04 | rs73246579 | 0.0001879 |
| *FAM21A* | 10 | 83 | PD | CD4NC | -0.526 | 1.448E-04 | rs7149844 | 4.84E-05 |
| *RP11_119F19_2* | 10 | 110 | PD | CD8ET | -0.490 | 3.837E-03 | rs8047876 | 0.0001835 |
| *MGMT* | 10 | 1025 | PD | BMem | -0.342 | 1.493E-03 | rs78966246 | 8.53E-05 |
| *IFITM2* | 11 | 15 | CD | CD4ET | 0.773 | 7.214E-05 | rs543379 | 4.96E-07 |
| *CD81* | 11 | 74 | PD | BIN | -0.639 | 2.650E-03 | rs4406421 | 0.0001934 |

| | | | | | | | | |
|---|---|---|---|---|---|---|---|---|
| *RPL27A* | 11 | 142 | PD | BIN | -0.360 | 3.210E-03 | rs4299314 | 6.52E-05 |
| *FNBP4* | 11 | 172 | AD | BMem | -0.924 | 3.324E-04 | rs1928464 | 1.44E-07 |
| *FNBP4* | 11 | 180 | AD | CD8ET | -0.753 | 2.183E-03 | rs1928464 | 1.44E-07 |
| *FNBP4* | 11 | 143 | AD | CD8NC | -0.760 | 2.029E-03 | rs1928464 | 1.44E-07 |
| *TMEM258* | 11 | 34 | CD | CD4ET | 1.000 | 8.847E-06 | rs102275 | 2.71E-07 |
| *FADS2* | 11 | 9 | CD | CD4NC | 1.000 | 7.141E-08 | rs102275 | 2.71E-07 |
| *SLC2A14* | 12 | 92 | PD | CD8ET | 0.255 | 3.044E-04 | rs4758263 | 0.0005911 |
| *RP11_428G5_5* | 12 | 103 | PD | BMem | 0.582 | 3.445E-03 | rs4106072 | 6.62E-05 |
| *CLLU1OS* | 12 | 82 | SCZ | BIN | 0.625 | 1.885E-03 | rs78822921 | 2.44E-05 |
| *RPL36AL* | 14 | 122 | PD | BIN | -0.238 | 3.246E-03 | rs982983 | 0.0003156 |
| *RPL36AL* | 14 | 129 | PD | CD4ET | -0.226 | 2.423E-03 | rs982983 | 0.0003156 |
| *RPL36AL* | 14 | 236 | PD | CD8ET | -0.264 | 4.253E-04 | rs982983 | 0.0003156 |
| *RPL36AL* | 14 | 212 | PD | CD8NC | -0.281 | 1.437E-04 | rs982983 | 0.0003156 |
| *KTN1* | 14 | 574 | PD | CD4NC | 0.454 | 1.064E-03 | rs12880964 | 0.0002186 |
| *KTN1* | 14 | 137 | PD | CD8ET | 0.615 | 2.133E-04 | rs12880964 | 0.0002186 |
| *SLC25A29* | 14 | 118 | PD | CD8ET | 0.225 | 1.725E-03 | rs1563783 | 3.00E-05 |
| *FAM98B* | 15 | 21 | SCZ | CD4NC | -0.735 | 2.245E-03 | rs478606 | 9.35E-08 |
| *NME4* | 16 | 147 | PD | BMem | -0.275 | 3.854E-03 | rs16990597 | 8.93E-05 |
| *TUFM* | 16 | 115 | PD | CD4ET | -0.349 | 3.736E-03 | rs28723531 | 7.90E-05 |
| *TUFM* | 16 | 122 | PD | CD8ET | -0.346 | 2.663E-03 | rs28723531 | 7.90E-05 |
| *CES1* | 16 | 47 | PD | CD4NC | -0.376 | 2.105E-03 | rs12880964 | 0.0002186 |
| *CES1* | 16 | 74 | PD | CD8ET | -0.339 | 3.037E-04 | rs12880964 | 0.0002186 |
| *CES1* | 16 | 58 | PD | CD8NC | -0.528 | 2.104E-03 | rs12880964 | 0.0002186 |
| *CES1* | 16 | 105 | PD | MonoC | -0.301 | 1.671E-03 | rs12880964 | 0.0002186 |
| *NUTF2* | 16 | 71 | SCZ | CD8NC | -1.000 | 9.134E-05 | rs7195605 | 2.03E-06 |
| *ELP5* | 17 | 133 | PD | CD4NC | -0.219 | 3.284E-03 | rs74662146 | 0.0002202 |
| *SPECC1* | 17 | 189 | SCZ | CD8ET | 0.737 | 1.911E-03 | rs8137258 | 9.87E-07 |
| *ORMDL3* | 17 | 241 | PD | BIN | -0.380 | 5.872E-06 | rs12147950 | 9.46E-06 |

| | | | | | | | | |
|---|---|---|---|---|---|---|---|---|
| *ORMDL3* | 17 | 189 | PD | BMem | -0.332 | 2.136E-04 | rs12147950 | 9.46E-06 |
| *ORMDL3* | 17 | 277 | PD | CD8ET | -0.204 | 3.444E-03 | rs12147950 | 9.46E-06 |
| *GSDMA* | 17 | 177 | PD | CD4NC | -0.474 | 4.691E-04 | rs1430035 | 1.06E-05 |
| *ST6GALNAC1* | 17 | 62 | PD | CD4ET | 0.855 | 4.628E-05 | rs146654644 | 5.39E-05 |
| *PGS1* | 17 | 95 | ALS | CD4NC | -0.761 | 1.223E-03 | rs12948394 | 1.81E-06 |
| *C17orf89* | 17 | 80 | PD | CD8NC | -0.426 | 5.697E-04 | rs181669976 | 0.00125 |
| *BSG* | 19 | 4 | PD | MonoC | -0.907 | 1.858E-04 | rs16990597 | 8.93E-05 |
| *AKAP8* | 19 | 88 | CD | CD4NC | -0.826 | 5.315E-04 | rs12459348 | 5.18E-07 |
| *AKAP8* | 19 | 80 | CD | CD8NC | -0.987 | 3.763E-04 | rs12459348 | 5.18E-07 |
| *SSBP4* | 19 | 127 | PD | CD8NC | 0.633 | 1.774E-03 | rs189987546 | 6.69E-05 |
| *PTGIR* | 19 | 133 | UC | CD4NC | -0.705 | 1.589E-05 | rs11667255 | 2.27E-07 |
| *PTGIR* | 19 | 29 | UC | CD8NC | -0.976 | 3.218E-05 | rs11667255 | 2.27E-07 |
| *GNG8* | 19 | 8 | UC | CD4NC | -1.000 | 7.313E-06 | rs11667255 | 2.27E-07 |
| *RPS5* | 19 | 112 | PD | BIN | 0.242 | 3.406E-05 | rs180724111 | 0.0003642 |
| *RPS5* | 19 | 87 | PD | CD8ET | 0.219 | 2.166E-03 | rs180724111 | 0.0003642 |
| *EDN3* | 20 | 14 | CD | CD8NC | -1.000 | 7.417E-04 | rs259958 | 3.01E-07 |
| *HAR1A* | 20 | 25 | PD | CD8ET | -0.398 | 1.814E-03 | rs149556186 | 0.0001065 |
| *PPDPF* | 20 | 30 | PD | BIN | -0.254 | 3.323E-03 | rs17303726 | 2.34E-05 |
| *PPDPF* | 20 | 42 | PD | CD8ET | -0.264 | 6.226E-04 | rs17303726 | 2.34E-05 |
| *PPDPF* | 20 | 36 | PD | CD8NC | -0.250 | 1.057E-03 | rs17303726 | 2.34E-05 |
| *SAMSN1* | 21 | 496 | UC | CD8ET | -0.584 | 1.795E-04 | rs2178936 | 5.04E-07 |
| *YBEY* | 21 | 431 | PD | MonoC | 0.448 | 2.053E-04 | rs35270498 | 5.72E-05 |
| *SLC25A1* | 22 | 87 | PD | CD8ET | 0.466 | 1.913E-03 | rs1206540 | 0.0002462 |
| *LGALS2* | 22 | 69 | PD | MonoC | 0.411 | 1.232E-03 | rs12147950 | 9.46E-06 |
| *RRP7A* | 22 | 65 | PD | CD8ET | -0.377 | 2.145E-03 | rs9914875 | 0.0003669 |
| *ARSA* | 22 | 30 | PD | CD8ET | 0.447 | 5.099E-05 | rs12443147 | 1.33E-05 |

AD = Alzheimer's disease; CD = Crohn's disease; MS = Multiple sclerosis; PD = Parkinson's disease; SCZ = Schizophrenia; UC = Ulcerative colitis; BMem = Memory B cells; CD4ET = CD4+ effector memory T cells; CD4NC = CD4+ naïve T cells; CD8ET = CD8+ effector memory T cells; CD8NC = CD8+ naïve T cells.
